# Exclusive Breastfeeding in the Follow-up of Preterm Infants: Challenges in Northern Brazil

**DOI:** 10.64898/2026.03.10.26347971

**Authors:** Ana Paula Del Tetto Zaccardi, Izabella Maria Pinheiro Palheta, Maria Luiza Del Tetto Zaccardi, Maiana Darwich Mendes Guerreiro

## Abstract

Exclusive breastfeeding (EBF) provides essential nutritional, immunological, and developmental benefits, particularly for preterm infants, who represent a vulnerable population. This study aimed to evaluate the duration of exclusive breastfeeding and investigate factors associated with early weaning among preterm infants followed in an outpatient clinic in Northern Brazil. A descriptive cross-sectional study was conducted through a retrospective review of medical records of preterm infants followed between November 2024 and April 2025. Neonatal and maternal variables were analyzed using descriptive statistics and Pearson’s correlation. A total of 69 preterm infants were included, with a mean gestational age of 33.79 weeks, plus or minus 2.97 weeks. Exclusive breastfeeding was observed in 41% of infants, with a mean duration of 3.84 months, plus or minus 1.99 months. The most frequent neonatal complications were jaundice (42%) and respiratory distress (41%). No statistically significant associations were identified between exclusive breastfeeding and the analyzed variables, although a positive trend was observed between the number of antenatal consultations and the duration of EBF (r = 0.5; p = 0.07). The findings indicate that exclusive breastfeeding among preterm infants remains below recommended levels, highlighting the importance of strengthening antenatal guidance and multiprofessional support to improve breastfeeding duration in this population.

## INTRODUCTION

Exclusive breastfeeding is defined by the Brazilian Society of Pediatrics (SBP) as the practice whereby an infant receives only human milk, whether directly from the breast, expressed, or obtained from another human source, without the introduction of any other liquids or solids.^1^ The World Health Organization (WHO), the Brazilian Ministry of Health, and the SBP recommend that exclusive breastfeeding should be maintained for the first six months of life, with complementary foods introduced thereafter whilst breastfeeding continues, preferably until at least two years of age.^1^

Human milk is a natural and biologically specialized food that provides all essential nutrients and a wide range of bioactive compounds during the period of exclusive breastfeeding. It contains proteins, lipids, immune cells, immunoglobulins, lactoferrin, lysozyme, and bifidum factor, all of which are fundamental to immune protection and infant development.^2,3^ Its composition is dynamic and varies according to the infant’s physiological needs, including gestational age at birth.^4,5^

This is particularly relevant for preterm infants, who represent a highly vulnerable population with increased nutritional, immunological, and neurological demands. Milk produced by mothers of preterm infants contains higher concentrations of proteins, lipids, free amino acids, sodium, copper, and zinc, as well as oligosaccharides and other components that support nutrient absorption, immune function, and neurodevelopment.^6,7^ Breastfeeding therefore represents a biologically tailored and protective intervention in this group.

The benefits of exclusive breastfeeding are well established for both mothers and infants. For mothers, it strengthens emotional bonding, supports hormonal regulation, contributes to the prevention of postnatal depression, and is associated with lower risks of hypertension, obesity, and breast cancer.^1,4^ For infants, it promotes appropriate growth and development, reduces the risk of diarrhea, respiratory infections, type 1 diabetes, and dyslipidemia, and supports orofacial and neurocognitive development.^1^ Among preterm infants, breastfeeding is also associated with lower rates of sepsis, necrotizing enterocolitis, retinopathy of prematurity, hospital admission, and adverse neurodevelopmental outcomes.^7^

By contrast, the early introduction of foods before six months offers no proven benefit and may be harmful, being associated with increased illness, reduced absorption of nutrients from breast milk, and diminished contraceptive effect of lactation for the mother.^1^ Weaning should therefore occur gradually, preferably between two and four years of age, with continued maternal support and guidance.^1,8^

Despite these recognized benefits, early weaning remains common among preterm infants. This is often related to difficulties with sucking, digestive immaturity, maternal–infant separation during hospitalization, and the emotional and physical burdens experienced by mothers. In Northern Brazil, early weaning has also been associated with socioeconomic, behavioral, and cultural factors, including household crowding, early use of dummies and feeding bottles, and replacement of breast milk with traditional regional foods such as açaí porridge and caribé.^10,11^

Although the benefits of breastfeeding in preterm populations are well documented, important gaps remain regarding the continuity of exclusive breastfeeding after hospital discharge and during outpatient follow-up, especially in socially and regionally vulnerable settings.^9,10,11^ In this context, multiprofessional follow-up from birth, including educational measures, psychological support, and structured breastfeeding promotion strategies, is essential.^9^ Based on this rationale, the present study aimed to evaluate the duration of exclusive breastfeeding and to investigate factors associated with early weaning among preterm infants followed in an outpatient setting.

## METHODS

### Ethical considerations

All participants included in this study were investigated in accordance with the principles of the Declaration of Helsinki and the Nuremberg Code, as well as Brazilian regulations governing research involving human participants (Law No. 14,874/2024). This study forms part of a broader project conducted at a preterm infant outpatient clinic and previously approved by the Research Ethics Committee as part of an umbrella project. The study was registered under CAAE number 79287824.5.0000.5169 and approved under opinion number 6,855,844.

### Study design, setting and sample

This was a descriptive cross-sectional clinical-epidemiological study with a quantitative approach, based on the review of medical records from patients followed at a preterm infant outpatient clinic in Northern Brazil between November 2024 and April 2025.

The study analyzed the prevalence of exclusive breastfeeding and the variables associated with it. Variables collected for the preterm infants included gestational age, length of hospital stay, sex, birth weight, nutrition, and most recent outpatient visit. Maternal variables included maternal age, antenatal care, primiparity, and mode of delivery.

### Data analysis

Data were organized into tables and graphs using Word and Excel and analyzed with BioEstat 5.0 software. Descriptive statistics included frequencies and measures of central tendency and dispersion. Inferential analyses were performed using Pearson’s correlation and multiple linear regression. Statistical significance was set at p ≤ 0.05.

## RESULTS

A total of 69 preterm newborns were included in the study. Mean gestational age was 33.79 ± 2.97 weeks, and mean length of hospital stay was 23.90 ± 21.19 days. Mothers had a mean age of 26.37 ± 5.88 years and attended a mean of 6.25 ± 2.96 antenatal consultations (Table 1).

**Table 1.** Characteristics of preterm infants attending an outpatient service.

| Variable | Mean | s.d. |
| --- | --- | --- |
| Gestational age | 33.79 | 2.97 |
| Length of hospital stay | 23.90 | 21.19 |
| Maternal age | 26.37 | 5.88 |
| Number of antenatal consultations | 6.25 | 2.96 |

Of the preterm newborns, 35 (51%) were male and 34 (49%) were female. Regarding birth weight, 3 infants (4%) were classified as extremely low birth weight, 12 (17%) as very low birth weight, 39 (57%) as low birth weight, and 15 (22%) as having normal birth weight.

With respect to feeding type, 28 children (41%) received exclusive breastfeeding (EBF), 15 (22%) were fed infant formula, 2 (3%) received complemented feeding, 23 (33%) received mixed feeding, and 1 child (1%) was consuming solid foods. Among those who received EBF, the mean duration was 3.84 ± 1.99 months. Mean overall breastfeeding duration was 9.77 ± 6.53 months (Table 2).

**Table 2.**
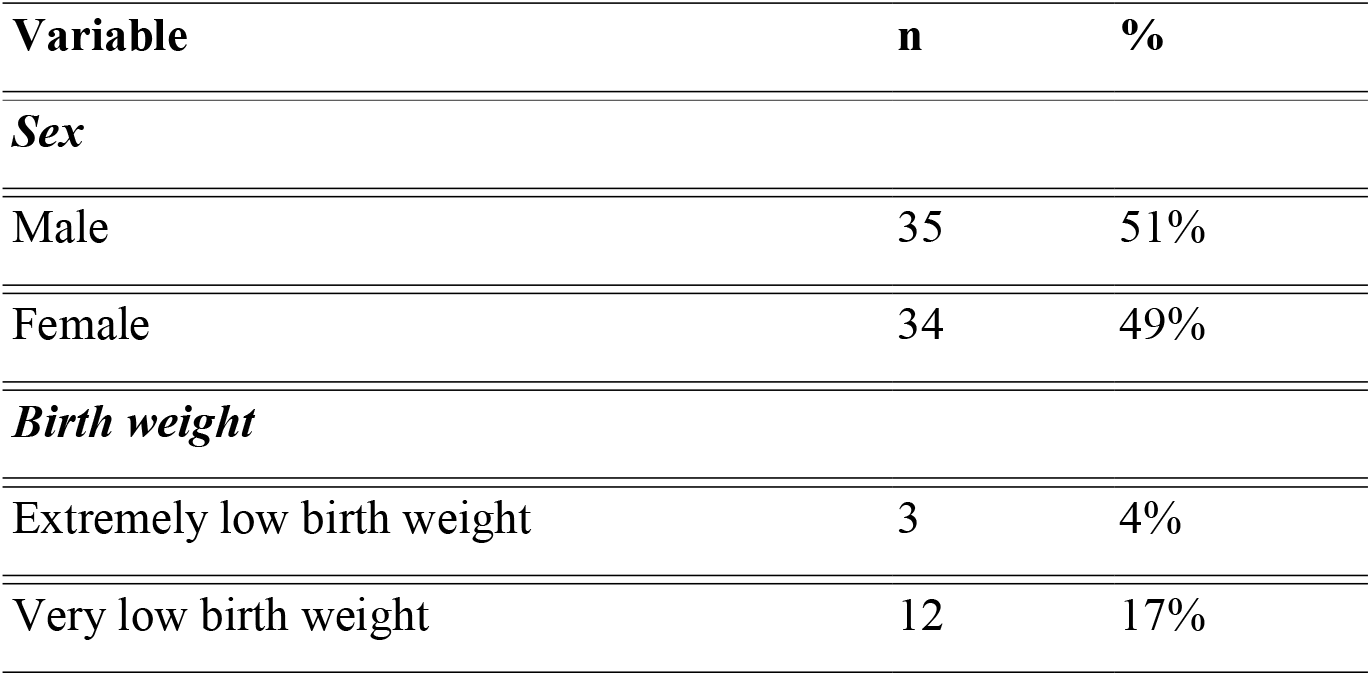
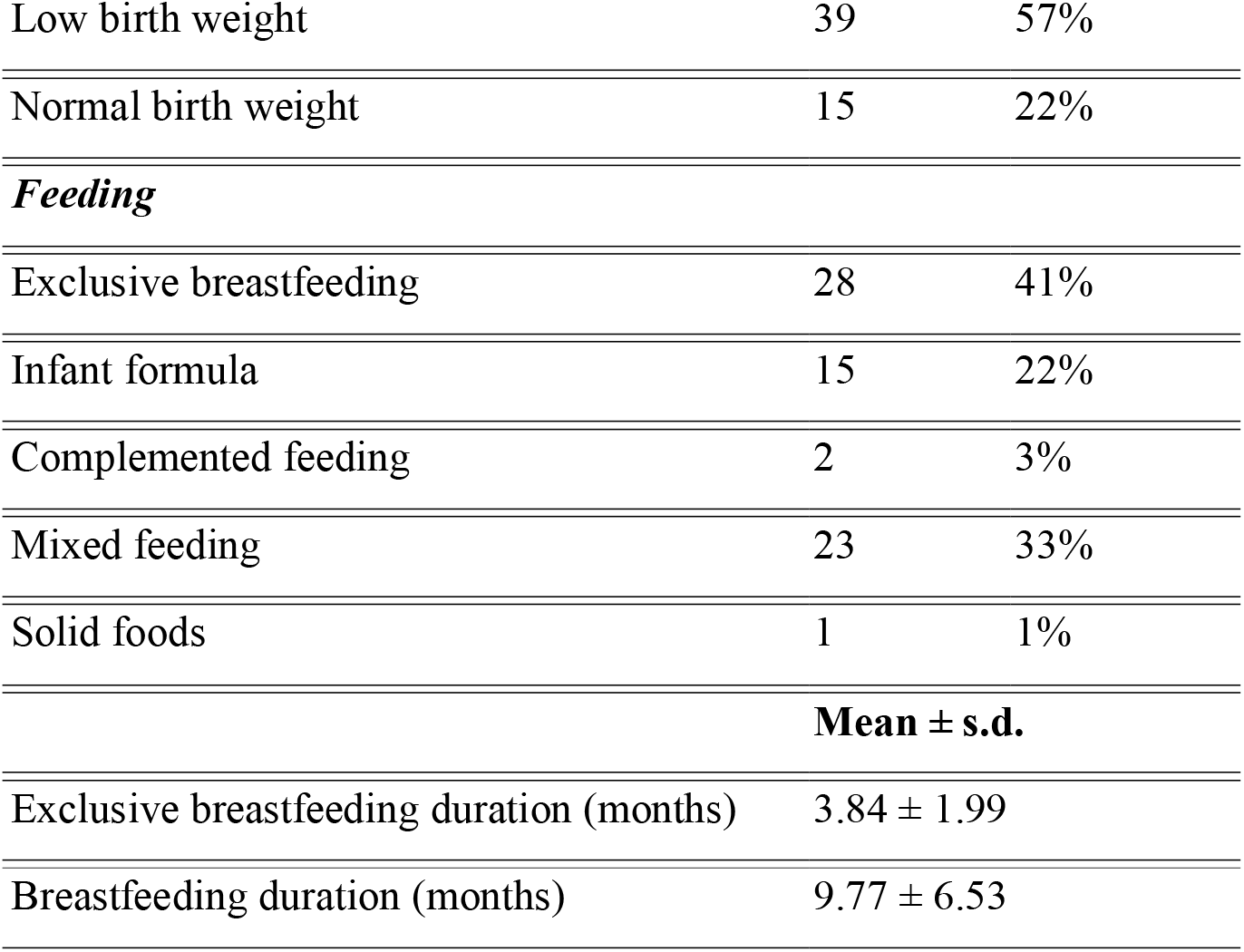
Sex, birth weight, and feeding profile of preterm infants attending an outpatient clinic.

With regard to maternal characteristics, most mothers were not primigravidae (n = 41; 60%), and most deliveries occurred by caesarean section (n = 44; 64%). In addition, 21 mothers (30%) had their most recent consultation recorded in 2024 (Table 3).

**Table 3.** Maternal characteristics of preterm infants attending an outpatient clinic.

| **Variable** | **n** | **%** |
| --- | --- | --- |
| ***Primigravida mother*** |  |  |
| Yes | 27 | 39% |
| No | 41 | 60% |
| Not reported | 1 | 1% |
| ***Mode of delivery*** |  |  |
| Caesarean section | 44 | 64% |
| Vaginal delivery | 23 | 33% |
| Not reported | 2 | 3% |
| ***Most recent consultation (loss to follow-up)*** |  |  |
| 2022 | 9 | 13% |
| 2023 | 10 | 14% |
| 2024 | 21 | 30% |
| 2025 | 16 | 23% |
| Not reported | 13 | 19% |

Regarding clinical manifestations, the most frequent findings were jaundice (n = 29; 42%) and respiratory distress (n = 28; 41%), followed by neonatal sepsis (n = 23; 33%) and hypoglycaemia (n = 11; 16%). Other conditions, such as resuscitation at birth (4%), cardiac abnormalities (3%), and electrolyte disturbances (3%), were less frequent. Neonatal infection, congenital syphilis, pneumothorax, traumatic brain injury (TBI), bradycardia, seizures, chylothorax, cyanosis, apnoea, and reflux were each reported in only 1% of cases (Table 4).

**Table 4.** Clinical manifestations among preterm infants attending an outpatient clinic.

| **Clinical manifestation** | **n** | **%** |
| --- | --- | --- |
| Jaundice | 29 | 42% |
| Respiratory distress | 28 | 41% |
| Resuscitation | 3 | 4% |
| Neonatal infection | 1 | 1% |
| Sepsis | 23 | 33% |
| Cardiac abnormalities | 2 | 3% |
| Electrolyte disturbance | 2 | 3% |
| Syphilis | 1 | 1% |
| Pneumothorax | 1 | 1% |
| Traumatic brain injury | 1 | 1% |
| Hypoglycaemia | 11 | 16% |
| Bradycardia | 1 | 1% |
| Seizure | 1 | 1% |
| Chylothorax | 1 | 1% |
| Cyanosis | 1 | 1% |
| Apnoea | 1 | 1% |
| Reflux | 1 | 1% |

Analysis of the association between exclusive breastfeeding and potentially influencing variables showed no statistically significant associations. However, a positive correlation was observed between the number of antenatal consultations and the duration of EBF (r = 0.5), suggesting a trend towards longer exclusive breastfeeding among mothers who attended more consultations. Although this trend was noteworthy, it did not reach statistical significance (p = 0.07) (Table 5).

**Table 5.** Association between exclusive breastfeeding and variables that may influence it.

| **Variable** | **r (Pearson)** | **p-value** |
| --- | --- | --- |
| Gestational age | 0.4 | 0.10 |

| Variable | r (Pearson) | p-value |
| --- | --- | --- |
| Birth weight | -0.0 | 0.75 |
| Primigravida mother | -0.2 | 0.45 |
| Maternal age | -0.1 | 0.52 |
| Number of antenatal consultations | 0.5 | 0.07 |
| Mode of delivery | 0.2 | 0.41 |
| Length of hospital stay | 0.2 | 0.45 |

Pearson's correlation test, $p \leq 0.05$ . *r*: Pearson's correlation coefficient.

Finally, multiple linear regression was performed to investigate the association between exclusive breastfeeding and the clinical complications observed in preterm children. No statistically significant relationship was identified among the variables analysed (F = 1.38; p = 0.43).

## DISCUSSION

The present study identified an exclusive breastfeeding (EBF) prevalence of 41% among preterm infants followed at an outpatient clinic in Northern Brazil, from admission to the clinic to a duration of less than four months. This finding differs from data reported in the 2000s, when an increase in breastfeeding prevalence was observed, particularly in the Northern Region of Brazil, with rates of 62% among children aged up to six months and 44% among those older than six months.^10^ Although these figures are relevant, they remain below the recommendation of the World Health Organization (WHO), which advocates the maintenance of EBF until six months of age.

This result is consistent with other Brazilian studies showing difficulties in maintaining exclusive breastfeeding among preterm infants. Many of these newborns require admission to neonatal intensive care units, experience early separation from their mothers whilst still fasting, and consequently depend on human milk banks. For example, an integrative review conducted in 2022 involving newborns with a gestational age of less than 37 weeks reported a median breastfeeding duration of five months. These findings may be explained primarily by physiological immaturity and by clinical conditions requiring prolonged hospitalisation.^9^

Analysis of neonatal characteristics showed a mean gestational age of 33.79 weeks, a mean hospital stay of approximately 24 days, and a high prevalence of clinical complications, particularly jaundice, respiratory distress, and sepsis. These factors have already been described in the literature as potential risks for early interruption of exclusive breastfeeding. In agreement with this, a study published in 2021 in the *Revista Baiana de Saúde Pública*, which investigated the nutritional status of preterm infants in an intensive care setting, found that 47% of infants were discharged on mixed feeding, thereby compromising the maintenance of EBF until the sixth month of life.^7^

With regard to maternal characteristics, most mothers had a mean age of 26 years, 60% were multiparous, and the average number of antenatal consultations was considered adequate. These data suggest that good-quality antenatal follow-up may act as a protective factor in the maintenance of exclusive breastfeeding. This interpretation is consistent with findings from the Maternal and Child Birth Cohort of Acre, Northern Brazil (MINA-Brazil), in which interruption of EBF before six months of age was more frequent among children born to primiparous mothers and among those exposed early to prelacteal feeds and dummy use, factors often associated with insufficient guidance during antenatal care, in addition to episodes of diarrhoea early in life.^11^

Other studies have likewise highlighted health education during pregnancy as a determining factor in breastfeeding success. A review conducted by H.S.M. Francislady, which investigated factors associated with the maintenance and interruption of EBF, found that 80% of participants had received guidance from health professionals during antenatal care and that 90% were aware of the benefits of breast milk. In that study, the prevalence of EBF between 7 and 15 days after hospital discharge was 66.4%.^8^ These results reinforce the importance of prenatal counselling and structured support as potentially modifiable factors in breastfeeding outcomes.

In the specific context of the state of Pará, although breastfeeding indicators appear to have improved over time, they remain below WHO recommendations. This pattern may be partly explained by the vast territorial dimension of the state, which poses challenges to the dissemination of information and to the provision of appropriate breastfeeding guidance, particularly for families of preterm newborns. In addition, there is still a shortage of robust studies examining comprehensively the factors that facilitate or hinder breastfeeding in this setting, which limits the design of more effective and context-sensitive intervention strategies.

Taken together, the present findings suggest that, although no statistically significant associations were identified, neonatal, maternal, and healthcare-related factors may still exert an important practical and clinical influence on the maintenance of exclusive breastfeeding. The absence of statistically significant results may reflect the relatively low prevalence of preterm infants receiving EBF in this outpatient population, as well as the fact that the sample was drawn from a single specialised medical centre. These aspects may have reduced the statistical power of the analyses and limited the detection of associations of smaller magnitude.

Nevertheless, the study contributes relevant evidence from a region where data on breastfeeding among preterm infants remain limited. By documenting both the prevalence of EBF and the clinical and maternal profile of this population, the findings help to clarify the local context of breastfeeding maintenance after hospital discharge. From a clinical and public health perspective, these results support the need for continuous multiprofessional follow-up, strengthening of antenatal and postnatal guidance, and targeted strategies to support mothers of preterm infants, especially in socially and geographically vulnerable settings.^7–11^

## CONCLUSION

The present study highlights the challenges involved in maintaining exclusive breastfeeding (EBF) among preterm infants, particularly in the context of physiological immaturity and clinical complications requiring prolonged hospitalisation. Although the prevalence observed was lower than that recommended by the WHO, the findings reinforce the importance of high-quality antenatal care and continuous support strategies for mothers, including guidance, encouragement of early breastfeeding, and multiprofessional assistance.

Accordingly, this study contributes to a better understanding of the barriers to EBF in this population and points to the need for targeted policies and interventions capable of increasing both the duration of and adherence to exclusive breastfeeding, thereby promoting nutritional, immunological, and developmental benefits for preterm newborns.

## Data Availability

All data produced in the present study are available upon reasonable request to the authors

https://doi.org/10.20396/san.v28i00.8658349

https://doi.org/10.33448/rsd-v11i2.25301

https://doi.org/10.11606/s1518-8787.2023057005563

